# Long, thin transmission chains of Severe Acute Respiratory Syndrome Coronavirus (SARS-CoV-2) may go undetected for several weeks at low to moderate reproductive numbers: Implications for containment and elimination strategy

**DOI:** 10.1101/2020.09.04.20187948

**Authors:** Gerry F. Killeen

## Abstract

Especially at low to moderate reproductive numbers, the generally mild, non-specific symptomology of Severe Acute Respiratory Syndrome Coronavirus (SARS-CoV-2) allows long, thin transmission chains to go undetected by passive surveillance over several weeks. This phenomenon has important implications: (1) Surveillance becomes less sensitive and reliable as an indicator of freedom from infection at the low reproductive numbers required to achieve elimination end points, passive surveillance systems may need to document an absence of new cases for at least a month to establish certainty of elimination. (2) Reproductive numbers should be kept as low as possible throughout such follow up periods without confirmed cases, to ensure such long, thin, undetected transmission chains all collapse before restrictions are eased and reproduction numbers are allowed to rebound. (3) While contact tracing systems may be highly effective when applied to large clusters in foci of elevated transmission where wide, rapidly expanding transmission chains are detected within two viral generations, large fractions of community transmission occurring through thinner, more extended transmission chains at lower reproductive numbers are often be too long to trace retrospectively and will be underrepresented in surveillance data. (4) Wherever surveillance systems are weak and/or younger age groups with lower rates of overt symptoms dominate transmission, containment effectiveness of contact tracing and isolation may be more severely limited, even at the higher reproduction numbers associated with larger outbreaks. While, contact tracing and isolation will remain vital for at least partially containing larger outbreaks, containment and elimination of SARS-CoV-2 will have to rely primarily upon the more burdensome and presumptive population-wide prevention measures that have proven so effective thus far against community transmission. Furthermore, these will have to be sustained at a much more stringent level and for longer periods after the last detected case than was necessary for SARS-CoV-1.

## Comparing and contrasting the pandemic trajectories of Severe Acute Respiratory Syndrome Coronavirus 1 and 2

As the world continues to struggle with the ongoing global pandemic of Severe Acute Respiratory Syndrome Coronavirus 2 (SARS-CoV-2),^1^ it is essential to contrast its distinctive epidemiological characteristics with those of Severe Acute Respiratory Syndrome Coronavirus 1 (SARS-CoV-1) so we can understand why these two pathogens have followed such dramatically different epidemic trajectories.^2^ SARS-CoV-1 was first identified as an emerging human pathogen in late 2002 and rapidly spread to 29 countries^3^ for similar reasons to those underpinning the rapid spread of SARS-CoV-2 across the globe earlier this year but with a starkly different endpoint:

> *The relatively prolonged incubation period allowed asymptomatic air travellers to spread the disease globally and resulted in more than 8000 cases in 2003*.^4^

Not only did SARS-CoV-1 spread internationally just as rapidly as SARS-CoV-2,^2,3^ it was also similarly difficult to distinguish from other common causes of illness, especially early in the onset of infection:

> *No individual symptom or cluster of symptoms has proved to be specific for a diagnosis of SARS. Although fever is the most frequently reported symptom, it is sometimes absent on initial measurement*^5^

Despite its similarly rapid spread, similarly non-specific symptomology and higher fatality rate, the epidemic curve of SARS-CoV-1 was rapidly crushed^6^ and then terminated with only 774 deaths occurring before the main outbreak ended in July 2003.^3^ While SARS-CoV-1 probably persists as a potential zoonotic threat in its original animal reservoir, human-to-human transmission of this virus may be considered eradicated because it no human case has been documented since four minor, brief, subsequent outbreaks in 2004.^3^

So why is it that SARS1 was so rapidly and decisively eliminated or eradicated while the SARS2 pandemic has spiralled so wildly out of control internationally? More worryingly, why has SARS2 proven so difficult to eliminate locally, even for countries that successfully shrank their national epidemics down to a handful of residual cases?

While pre-symptomatic transmission is considered a key feature of SARS-CoV-2 epidemiology and one that facilitates its spread into new contexts^7^, this phenomenon alone does not necessarily preclude effective contact tracing and isolation but rather exacerbates the requirement to do so promptly and with greater retrospective reach over several days^2^. The biggest challenge with respect to containment of SARS-CoV-2 appears to be extensive community transmission arising from the common occurrence of infections causing only mild, non-specific symptoms if any, so that they evade detection through passive, facility-based surveillance of self-reporting individuals seeking testing and care^2,8-11^.

Here I develop and apply a simple, deterministic mathematical model to outline how the lower population-wide mean clinical severity of SARS-CoV-2 infections allows long, thin transmission chains to extend for several weeks without being detected through conventional, passive surveillance of self-reporting symptomatic cases. I then explain how this phenomenon necessitates stringent preventative measures to be sustained for at least a month after the last confirmed case before epidemics can be confidently considered eliminated, may facilitate undetected spread through focal transmission hubs, and may strongly bias epidemiological data towards more readily traceable cases in larger clusters wherever higher local reproductive numbers occur. I also outline how such extended covert transmission chains render contact tracing and isolation far less effective as a containment strategy than for SARS-CoV-2, especially at the low-to-moderate reproductive numbers representative of community transmission following effective suppression.

## Long, thin chains of asymptomatic SARS-CoV-2 transmission can go undetected for several weeks at low to moderate reproductive numbers

Although the clinical manifestations of viral infections are distributed along a continuum spectrum of severity, for simplicity, new infections are assumed here to result in one of three symptomological category outcomes, each associated with a distinct probability of self-reporting to clinical facilities or otherwise seeking testing at disease-specific testing centres: (1) The proportion of people infected who experience severe symptoms at some stage over the full course of their infections (*θ_s_*) and consequently all report for testing, (2) The proportion of infections that, sooner or later, result in some mild symptoms (*θ_m_*), sometimes referred to as *paucisymptomatic* infections, of which only a fraction self-report for testing (*ρ_m_*), and (3) the remaining proportion of infections that do not result in any obvious symptoms (*θ_a_* = 1 – *θ_s_* – *θ_m_*) and consequently do not self-report for testing. Based on the values assumed for these input parameters, the proportion of infected persons self-reporting for testing (*θ_r_*) may be calculated as:

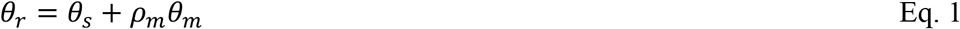

Allowing for the imperfect sensitivity of currently available diagnostic tests for viral genetic material, only a proportion of all self-reported active infections will be successfully confirmed when tested (*ρ_c_*), so the proportion of infections successfully tested and confirmed by routine symptom-based passive surveillance may be calculated thus:

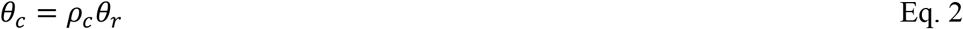

For SARS-CoV-1, these input parameters were set at *θ_s_* = 0.9, *θ_m_* = 0.05 and *θ_a_* = 0.05 based on previously reported low rates of mild or asymptomatic infections among a carefully monitored cohort of healthcare workers^12^. For SARS-CoV-2, these values were set at *θ_s_* = 0.1, *θ_m_* = 0.4 and *θ_a_* = 0.5, based on previous literature review^8^. For both coronaviruses, approximately half of all mildly symptomatic infections were assumed to self-report for testing (*ρ_m_* = 0.5) based on literature review relating to SARS-CoV-2^8^ and test sensitivity of 70% was assumed in both cases (*ρ_c_* = 0.7) based on performance of current SARS-CoV-2 tests^13^, so that the contrasting simulations for both viruses can be considered on an “all other things being equal” basis. Based on these assumed input parameters, the derived overall probabilities of infection confirmation through routine surveillance are calculated as approximately 65% for SARS-CoV-1 and 21% for SARS-CoV-2 (*θ_c_* = 0.6475 and 0.21, respectively).

The mean number of secondary cases arising from the primary cases that seed an epidemic in a given context, circumstance and intervention scenario, is commonly known as the effective reproductive number (*R_e_*). Here, that baseline reproductive number specifies an intervention scenario lacking tracing and isolation of contacts for confirmed cases identified through routine, symptom-based passive surveillance at health facilities and testing centres. After a given number of complete viral generation (*g*) durations of approximately two weeks each, the relative size of the *F_1_* viral population compared to the original *F_0_* population that seeded the epidemic before interventions were introduced (*V_g_*) may be calculated as:

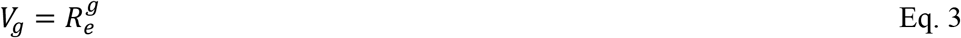

The overall probability that a transmission chain seeded by a single primary case remains unconfirmed after a given number of descendant secondary generations (*P_uc,g_*) may then be calculated as the same probability for the previous generation multiplied by the proportion of infections evading detection through routine surveillance (1 – *θ_c_*) to the power of the expected number of active infections derived from the primary infection by that generation:

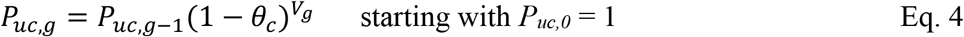

As illustrated in figure 1A, the median lengths of SARS-CoV-1 transmission chains up to the point where they are identified, tested and confirmed through routine passive, symptom-based surveillance are consistently short. Even when effective reproduction numbers drop slightly below the threshold required to sustain them (*R_e_*< 1.0), SARS-CoV-1 transmission chains rarely extend beyond 3 secondary generations. Looking at the lower end of this spectrum of reproduction numbers, it should be possible to use contact tracing to even contain SARS-CoV-1 transmission chains that are unusually extended because they are nevertheless detected sufficiently early to remain narrow enough to trace comprehensively through a single generation (Figure 2A).

**Figure 1.**
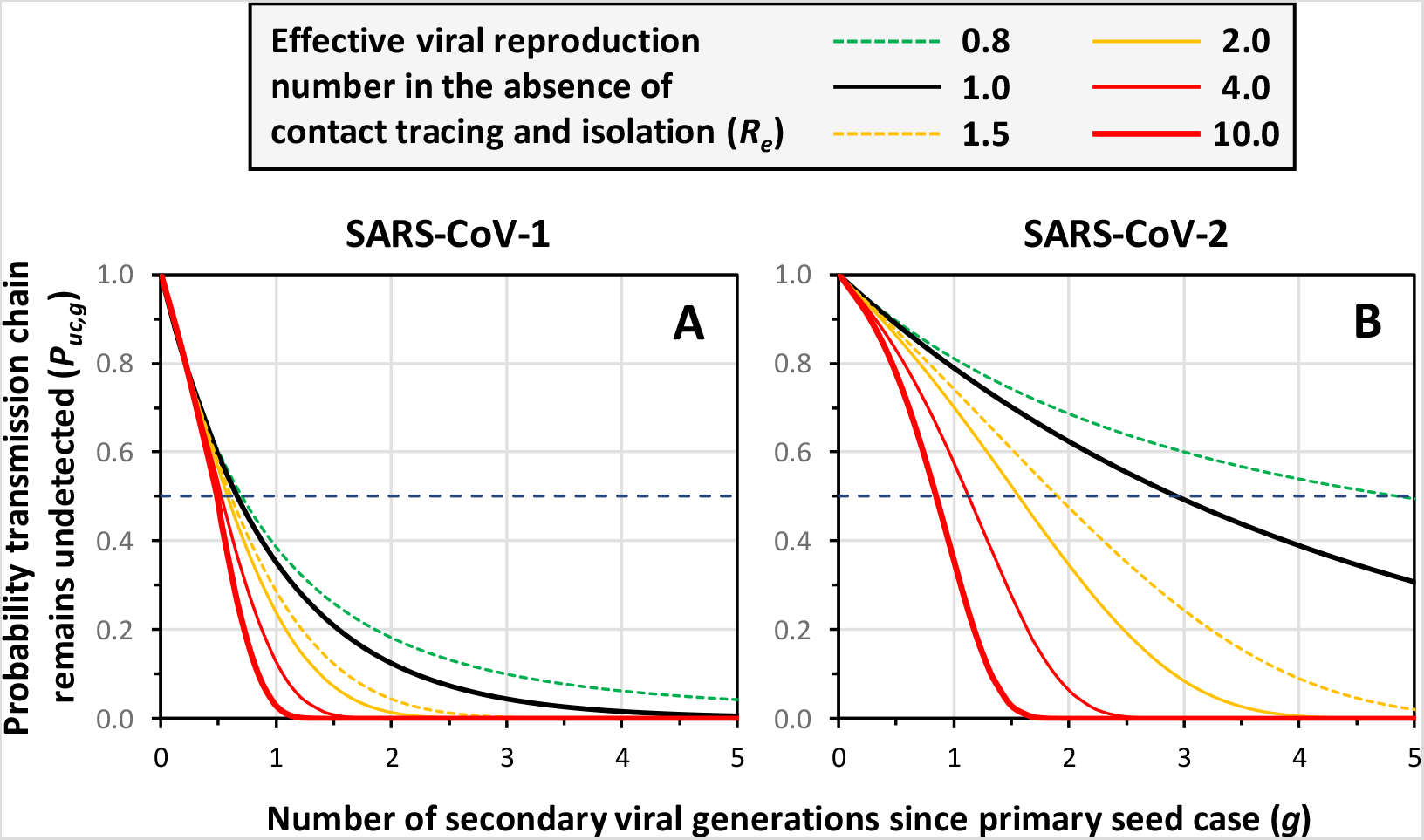
The declining probabilities over time, expressed in terms of full viral generation durations (*g*), each lasting approximately two weeks but overlapping so approximately one week apart for both SARS-CoV-1 and SARS-CoV-2,^2^ for transmission chains seeded by single individual primary cases remain unconfirmed by routine passive surveillance of self-reporting symptomatic cases (*P_uc,g_*), assuming a range of values for the effective reproductive number in the absence of any contact tracing and isolation intervention (*R_e_*). The horizontal dashed lines in both panels represents a probability of 50%, from which median chain length can be interpolated onto the horizontal axis.

**Figure 2.**
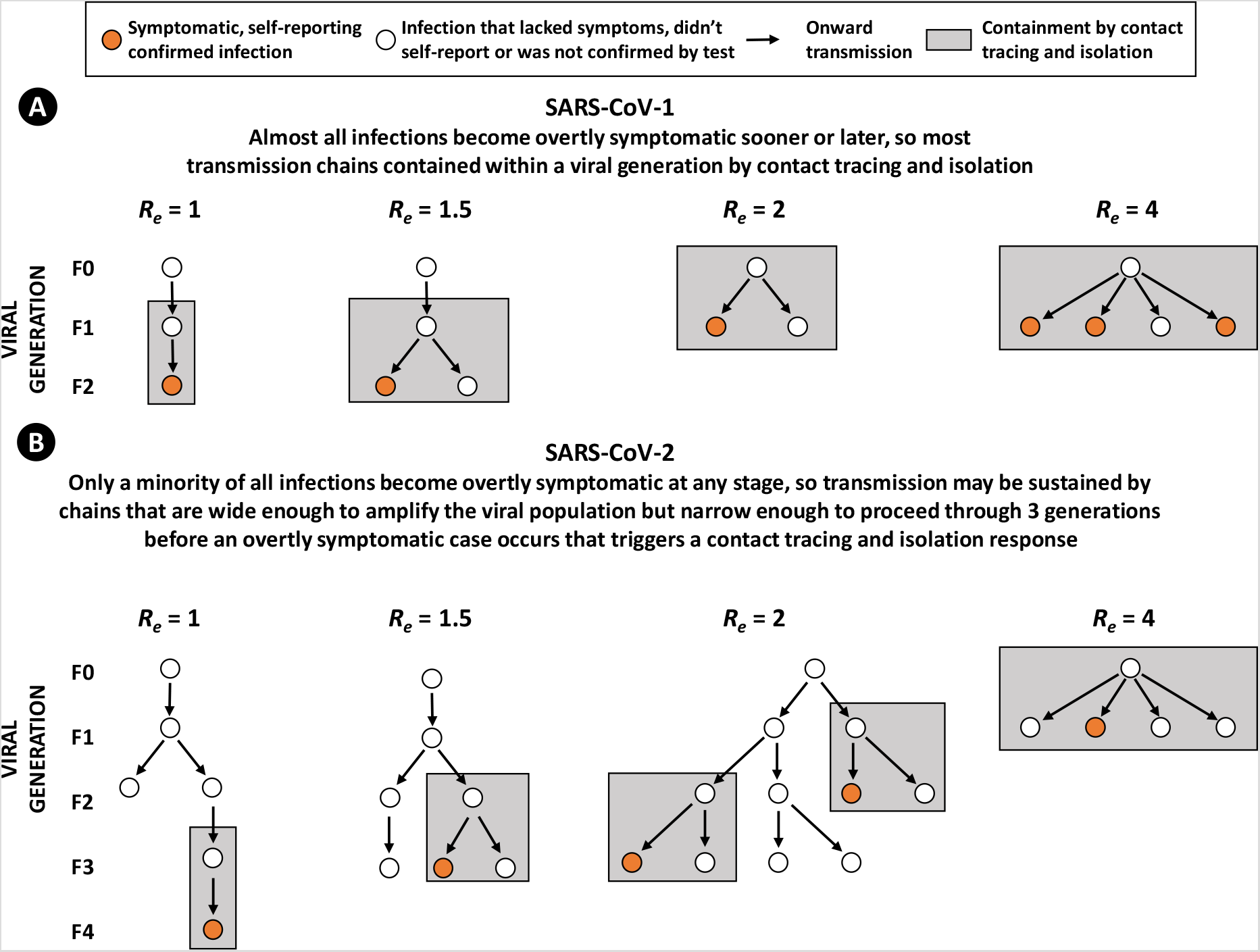
A schematic illustration of how (**A**) SARS-CoV-1 transmission chains may be consistently contained through effective contact tracing and isolation because the vast majority of infections result in clinical symptoms overt enough to motivate self-reporting to health facilities or testing centres, whereas (**B**) for SARS-CoV-2 only a small minority of infections cause sufficiently overt symptoms to motivate self-reporting, so transmission chains often extend too long before being detected for retrospective contact tracing and isolation to completely contain them.

At moderate effective reproductive numbers that are nevertheless comfortably above this minimal self-sustaining level and capable of seeding rapid exponential growth (*R_e_* = 1.5), it may be expected that almost three quarters (72%) of all transmission chains will be identified either at the point of the seeding primary infection or within a single secondary generation (Figure 1A) and 96% within 2 secondary generations. At the higher effective reproductive numbers that drive explosive outbreaks (*R_e_* ≥ 2.0), the vast majority (≥77%) of SARS-CoV-1 transmission chains should be detected and contained within a single secondary generation and essentially all (≥99%) within two secondary generations, even though a substantial minority of infections (35%) evade detection because of either the absence of overt symptoms or, more likely, false negative tests (Figure 1A). So regardless of effective reproductive number, essentially all SARS-CoV-1 transmission chains are expected to be identified and contained before any early branches have the opportunity to extend beyond the traceability limit of one subsequent generation (Figure 2A).

## Implications for surveillance and suppression of community transmission during the final stages of SARS-CoV-2 elimination programmes

Nevertheless, the phenomenon of long, thin transmission chains going undetected for so many generations through passive surveillance, even at reproductive numbers too low for those chains to sustain themselves indefinitely, has important implications for defining how long is required to be sure an epidemic has really ended. In stark contrast with SARS-CoV-1, even at a steady state reproductive number of 1.0 that yields essentially linear transmission chains on average, 39% of SARS-CoV-2 chains are expected to evade detection for up to 4 secondary generations (Figure 1B). Assuming a mean infection duration of two weeks, and a mean serial interval of one week for both coronaviruses^2^, 4 generations is equivalent to approximately one month.

The probability that a single transmission chain can sustain itself and remain unconfirmed (*P_suc,g_*) by evading detection through passive surveillance may be calculated as:

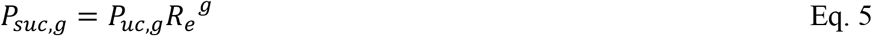

Looking specifically at the range of reproductive numbers below 1, at which elimination is achievable sooner or later, the contrast between SARS-CoV-1 and SARS-CoV-2 is stark in terms of this predictor of covert transmission chain persistence (Figure 3). At lower reproductive numbers, while most transmission chains will collapse before extending for so many generations, a small but important minority of SARS-CoV-2 chains that do survive are likely to remain undetected (Figure 3B and 3D) because they are so long and thin (Figure 2B). This has two important implications for the end-game of SARS-CoV-2 elimination programmes: First, surveillance becomes less sensitive and reliable as an indicator of freedom from infection at the low reproductive numbers required to achieve elimination end points, so even the best passive surveillance systems may need to document zero new cases for at least a full month to establish with reasonably satisfactory certainty (P ≤ 0.01) that elimination has been achieved and stringent restrictions may be eased (Figure 3B and D). Second, reproductive numbers should be kept as low as possible throughout such follow up periods with apparently no cases, to ensure such long, thin, undetected transmission chains all self-terminate before restrictions are eased and reproduction numbers are allowed to rebound (Figure 3B and D). In simple terms, restrictions effective enough to achieve *R_e_* ≤ 0.5 must be sustained throughout such a follow-up period to ensure satisfactory certainty of elimination, even after a full month (Figure 3B and 3D).

**Figure 3.**
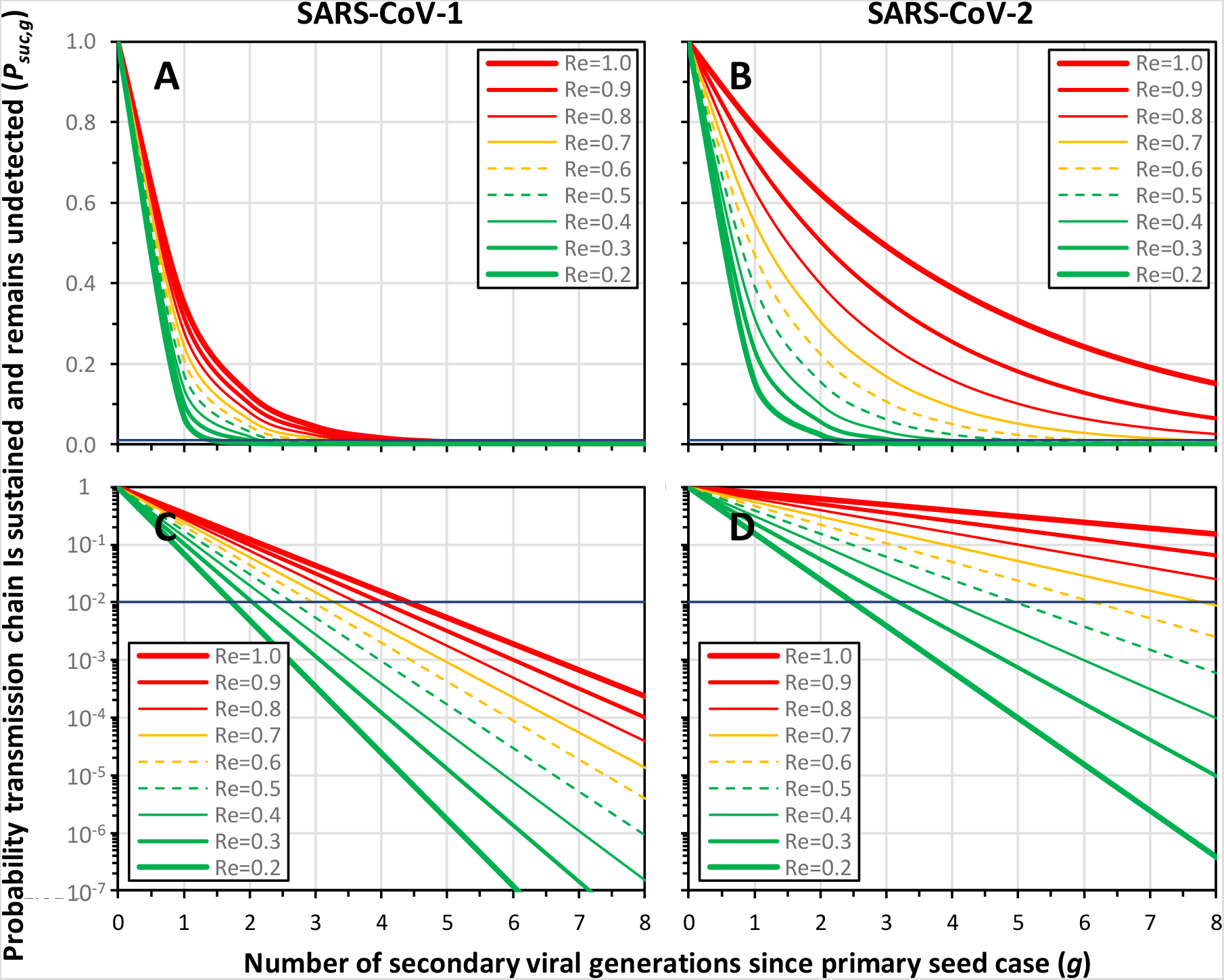
Predicted probabilities that individual transmission chains of SARS-CoV-1 (**A** and **C**) or SARS-CoV-2 (**B** and **D**) will both persistently sustain themselves and remain undetected and unconfirmed (*P_suc,g_*) after a given number of overlapping secondary generations (*g*) approximately one week apart^2^ by routine passive surveillance at reproductive numbers low enough to facilitate virus elimination. The horizontal lines in both panels represent a 1% probability threshold at which a period without any confirmed cases may be considered to reflect de facto elimination with a reasonably satisfactory degree of confidence (*P_suc,g_* = 0.01).

In contrast, for SARS-CoV-1 (Figure 3A and C), satisfactory certainty of elimination may be achieved within a month, even at the maximum reproductive number low enough to prevent epidemic resurgence (*R_e_* = 1.0). While a range of effective reproductive numbers have been simulated for both coronaviruses here to ensure direct comparability on the basis that all other things are equal, in reality the basic reproductive numbers of SARS-CoV-1 was substantially lower than for SARS-CoV-2,^2^ so this key difference in elimination requirements is likely to be even more important than figure 3 suggests.

## Implications for defining realistic expectations of contact tracing and isolation as a SARS-CoV-2 containment measure

Reproductive rates that are only moderately higher, but nevertheless sufficient to drive strong, steadily branching outbreaks of SARS-CoV-2 (*R_e_* = 1.5), can also generate remarkably extended transmission chains that may go unnoticed before an overtly symptomatic infection occurs, self-reports and is confirmed with laboratory diagnostics (Figure 1A). Indeed, such epidemic growth of 50% per generation results in transmission chains that are expected to usually branch once in the first two secondary generations but also to extend for three secondary generations or more before being detected in 24% of cases. As a result, branch points may commonly occur too long ago to be realistically traced retrospectively, so containment may be only partially effective (Figure 1B). While only 8.4% of transmission chains will span three secondary generations or more at a slightly higher effective *R_e_* of 2.0 (Figure 1B), this is counterbalanced by their faster rate of expansion and greater likelihood of branching too early to allow consistent tracing to single common ancestor infections by the time a confirmed index case is identified (Figure 2B).

Explosive epidemic growth at Re ≥ 4.0 shortens the duration over which SARS-CoV-2 transmission chains may go undetected (Figure 1B), because their greater width will usually yield an early confirmed index case in the first secondary generation (Figure 2B). Nevertheless, it is notable that 6% of such rapidly widening transmission chains are expected to extend to the F2 secondary generation before any symptomatic case is confirmed, at which point it would be too late to trace the initial branch point of the primary case (Figure 4). While such untraceable early branch points in extended, unnoticed transmission chains are rarer at such high reproductive numbers, their consequences are obviously greater simply because those escapee lineages amplify themselves so quickly (Figure 4).

**Figure 4.**
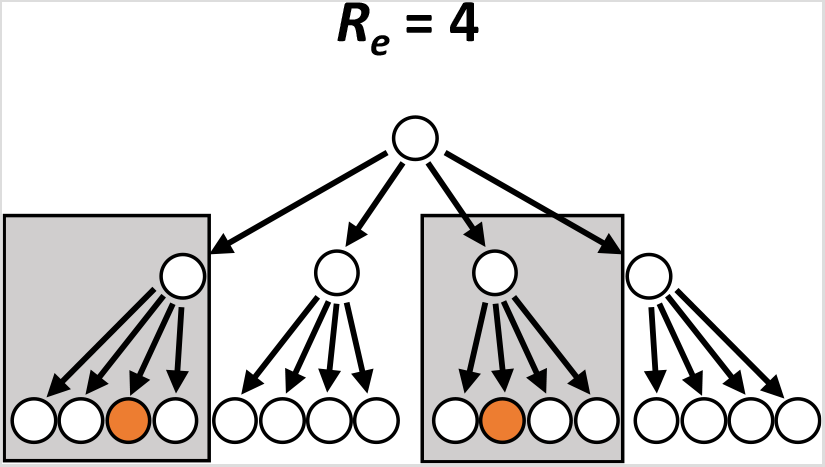
A schematic illustration of how wide, rapidly expanding transmission chains for SARS-CoV-2 can sometimes extend over three generations before being detected, at which point they most probably branched far too early for contact tracing and isolation to completely contain them.

The implications of this key interaction between the length of unnoticed transmission chains and their width may be explored numerically, first by calculating the probable fraction of transmission chains that evade retrospective tracing for each secondary generation (*P_uct,g_*):

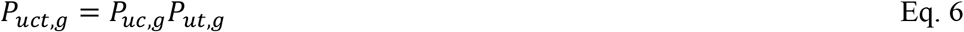

where

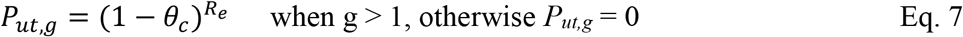

These biodemographic analyses of transmission chains indicate that extended chain lengths at low to moderate reproductive numbers (Figure 2B) may allow substantial numbers of viral lineages to escape from containment by contact tracing and isolation (Figure 5). For SARS-CoV-1, untraceable branches in transmission chains almost never occur, regardless of transmission potential (*R_e_*), simply because the overt and consistent symptomology of the disease always draws attention to itself, so chains are almost always identified at the stage of the primary infection or the first secondary generation (Figure 5A, C, E, G and I, plus Figure 2A for an intuitive explanation). For SARS-CoV-2, however, much longer chains can occur before detection with routine passive surveillance, especially at lower reproductive numbers (Figure 1), and this allows sufficient time for early branches of chain expansion to become untraceable before the first index case is confirmed (Figures 2A and 5B, D, F, H and J).

**Figure 5.**
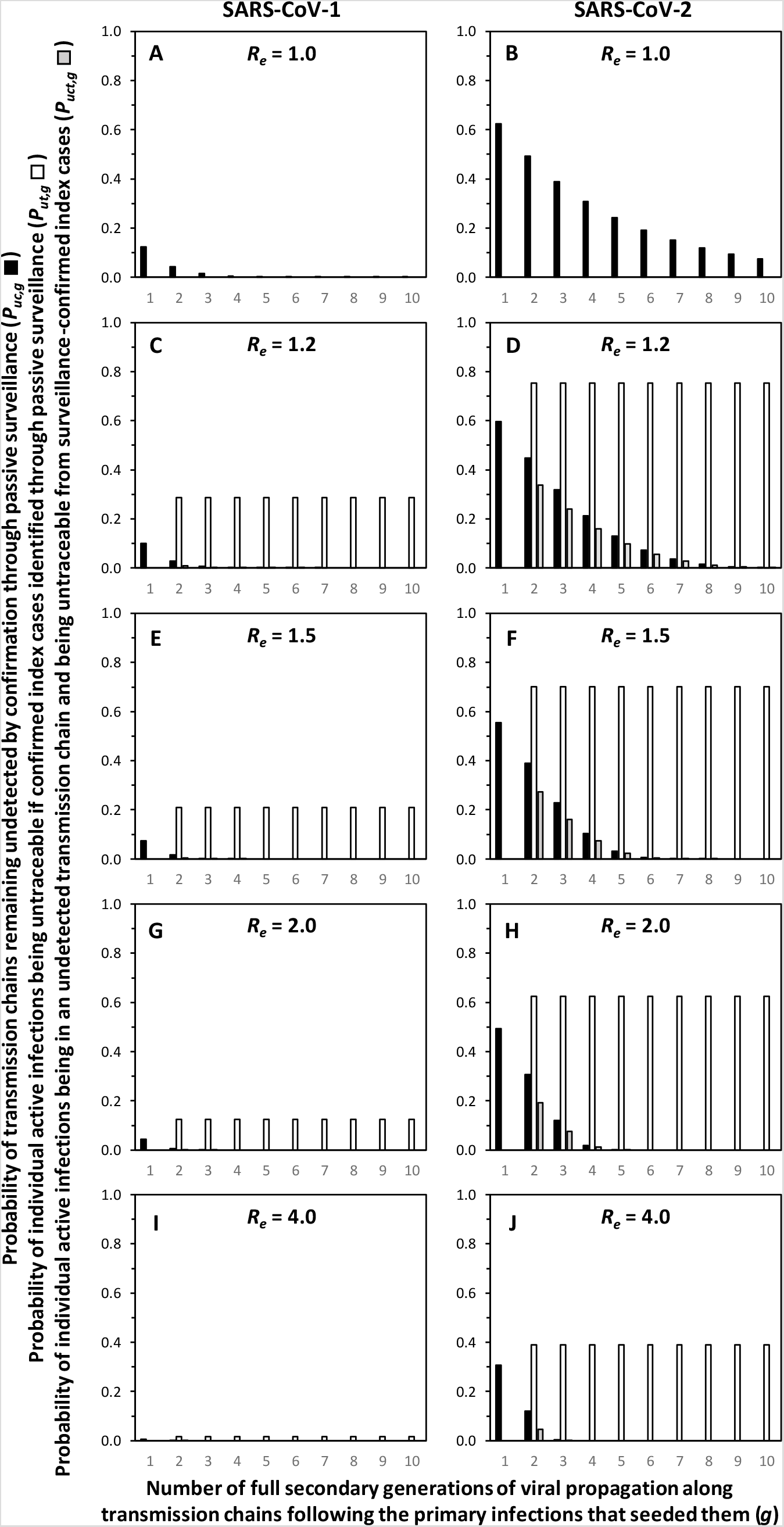
Predicted probabilities for each secondary viral generation that transmission chains of SARS-CoV-1 and SARS-CoV-2 remain undetected, that individual infections are untraceable because they originate from a common ancestor branch point more than two generations ago, and that untraceable infections arise from undetected transmission chains.

At least some degree of viral population expansion is required to allow some infections to evade containment through contact tracing because that requires at least occasionally sustained branching (Figure 2B). This intuitive descriptive rationale is reflected numerically in the lack of any predicted escapee infections in figure 5B despite the expectation that some of these essentially linear chains may extend for up to 10 generations before being detected through routine surveillance. However, while higher reproductive numbers obviously accelerate early branching and viral population expansion, this risk is counterbalanced by the fact that such wider chains will also be identified and confirmed earlier through routine surveillance (Figure 5B, D, F, H and J). Overall, the probability that SARS-CoV-2 transmission chains will yield untraceable offspring therefore peaks consistently in the F2 secondary generation and tails off rapidly in subsequent generations at higher reproductive numbers. At low to moderate reproduction numbers (1.0 < Re ≤ 2), modest rates of expansion are allowed to proceed through longer transmission chains with much higher overall probabilities of producing untraceable offspring infections (Figure 5D, F and H).

However, these untraceability probabilities alone underrepresent the influence of reproductive numbers because the wider transmission chains in later secondary generations represent larger viral populations, from which any given proportion of untraceable infections will amount to a greater number of escapees overall. The expected mean number of viral infections that cannot be contained by contact tracing and isolation because they branched off two generations or weeks ago (*V_uct,g_*) may be calculated for each secondary generation as the product of the probability of infections being both unconfirmed and untraceable (*P_uct,g_*) and the otherwise expected number of viral offspring descended from the primary seed infection (*V_g_*):

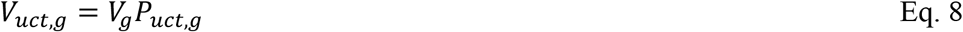

The expected mean number of infections arising from entire transmission chains over their full duration that are fundamentally untraceable, and will escape containment regardless of how effectively contact tracing and isolation is implemented because their nearest common ancestors with confirmed cases occurred two or more generations previously (*E*), can then be calculated by summing these probabilities per generation over the lifetimes of the transmission chains:

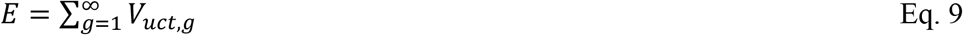

Similarly, the mean length of transmission chains, in generations, before they are detected through confirmed cases identified through routine passive surveillance (*G*) by summing the per-generation probabilities for chains remaining unconfirmed (*P_uc,g_*):

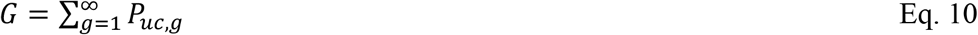

Figure 6 illustrates how the distribution of probabilities of transmission chains remaining undetected and then yielding untraceable escapee infections is modified by accounting for the fact that they are rapidly widened by higher reproductive numbers. At our default assumption for the proportion of infections detected and confirmed through passive surveillance (*θ_c_* = 0.21), high reproductive numbers so rapidly curtail the distribution of probabilities that transmission chains remain undetected (Figure 6A). Consequently, even the concomitantly greater population size of later generations is compensated for because so few chains ever get the chance for offshoot lineages that survive more than one generation beyond their common branch point (Figure 6B).

**Figure 6.**
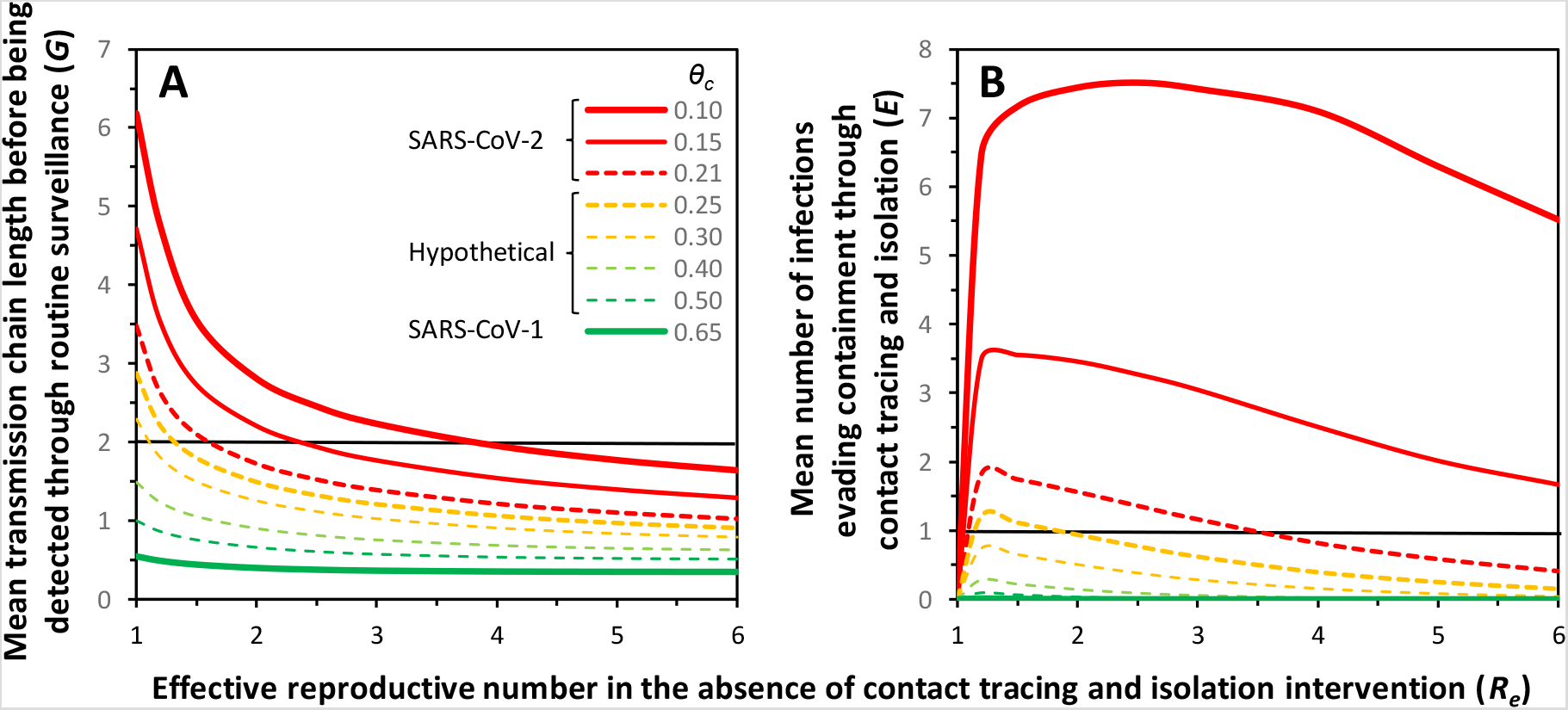
A comparison of the simulated potential for SARS-CoV-1 and SARS-CoV-2 and a series of otherwise identical hypothetical viral pathogens with intermediate rates of infection confirmation through routine surveillance (*θ_c_*) to evade containment through contact tracing and isolation. **A**: Predicted mean length of viral transmission chains up to the point at which they are detected through routine passive surveillance. **B**: Predicted mean number of untraceable infections arising from individual transmission chains. For SARS-CoV-2, a range of infection confirmation rates are assumed, to match either the approximate rates assumed for figures 1 to 5 based on previous literature review (*θ_c_* = 0.21)^8^, or similar to some even lower recent estimates (*θ_c_* = 0.15)^9-11^ that may be further attenuated in settings where surveillance systems are weaker and/or ongoing transmission is dominated by younger age groups who are less likely to become overtly symptomatic and self-report (*θ_c_* = 0.10). The horizontal line in panels **A** represents the minimum number of secondary generations, which approximates to weeks, that a transmission chain should extend without being detected for some of its earliest branch points from the primary generation to be considered untraceable. The horizontal line in panel **B** represents the minimum mean number of escapees per transmission chain required for those chains to robustly sustain themselves despite highly effective contact tracing and isolation because of early branch points that occurred too long ago to be traced once the first symptomatic case is tested and confirmed.

The highest rates at which transmission chains result in untraceable escapees occur at lower reproductive numbers which are just sufficient to sustain steady expansion of transmission chains and outbreaks in the absence of contact tracing and isolation. Crucially, the estimated mean number of untraceable escapees for SARS-CoV-2 approaches two per chain at reproductive numbers ranging up to about 2 and comfortably exceeds the threshold of 1 per transmission chain required for epidemic persistence up to effective reproductive numbers of 3 or more.

In stark contrast, SARS-CoV-1 is clearly grossly incapable of evading contact tracing over multiple generations (Figure 6), confirming the crucial importance of consistently overt symptoms of infection for enabling effective contact tracing and isolation rapidly enough to contain transmission chains before they become too extended (Figure 5). Simulations for a range of otherwise identical hypothetical viral pathogens with intermediate rates of detection through routine, symptom-driven, passive surveillance suggest that any substantive increase in detection rate above those typical of estimates for SARS-CoV-2 would close off this escape route for transmission chains with early branches (Figure 6). The infection confirmation rates assumed for SARS-CoV-2 in the simulations presented in figures 1, 3 and 5 appear to be just low enough for it to robustly evade containment with contact tracing and isolation (Figure 6), so more effective alternatives to passive, symptom-based surveillance could be considered as options for curtailing the progression of transmission chains before they become too long and branched to completely contain. Probably the most advanced and well-characterized operational models for active, population-wide surveillance of pathogens causing similarly vague, non-specific symptoms have been developed for malaria, HIV and tuberculosis in Africa and much could be learned from ongoing African initiatives to adapt these community-based approaches to SARS-CoV-2.^14^

The prediction that untraceability peaks at robust levels when reproductive numbers are low to moderate (Figure 6B) may be particularly useful for helping rationalize the recent experiences of many countries centres that have found their contact tracing systems to be highly effective when applied to large clusters but nevertheless leave large fractions of community transmission of untraceable, unexplained origin. In large clusters where local effective reproductive numbers are very high (*R_e_* estimates exceeding 10 have been reported for some settings like quarantined cruise ships^15^), almost all such wide transmission chains should be detected within a single secondary generation, and therefore traceable in principle over the week-long mean serial interval of SARS2-CoV-2.^2^ However, longer, narrower transmission chains that go unnoticed for several viral generations are probably typical of most community transmission events occurring outside of obvious large clusters, at the lower range of the reproductive numbers simulated in figure 6. Assuming the 80-20 rule holds true for SARS-CoV2 in the same way that it holds for every other known infectious disease,^16^ most people probably lived in social networks with reproductive numbers substantially below the reported range of estimated mean baseline reproductive numbers of 3 to 4 for this virus,^8^ even before any complementary pre-emptive, presumptive, population-wide behavioural interventions were rolled out across so many countries as the pandemic unfolded.

Such modest, but nevertheless self-sustaining reproductive numbers may therefore be the rule rather than the exception, especially in countries attempting to ease off the population-wide pre-emptive, presumptive hygiene measures and behavioural restrictions^1^ that are too socially burdensome and economically damaging to sustain indefinitely.^14,17^ After successfully crushing the curves^6^ of their initial epidemics, many countries have transitioned to more targeted, reactive end-game strategies relying more heavily on contact tracing and isolation^1^, similar to those that proved so successful against SARS-CoV-1.^2^ However, in the case of SARS-CoV-2, this approach may not be reasonably expected to play an important role in containing community transmission or for finishing off the last embers of transmission at the final stages of elimination programmes, at least until active, presumably community-based surveillance systems^14^ are developed that can consistently find otherwise covert transmission chains within two weeks. While these theoretical simulations are no more than that, they do paint a picture that strongly supports early perspectives shared by investigators with in-depth experience of both SARS-CoV-1^12^ and now SARS-CoV-2:

> *…many more patients with COVID-19 rather than SARS have mild symptoms that contribute to spread because these patients are often missed and not isolated*.^2^

Such an Achilles’ heel for contact tracing and isolation in settings with low-to-moderate transmission potential may help explain why so many countries have been seen their epidemics begin to accelerate again when attempting to use this containment option as a substitute for all the higher burden social distancing, hygiene and mask wearing interventions that helped them suppress their epidemics in the first place. So while development of improved diagnostics and surveillance platforms to push SARS-CoV-2 infection detection rates comfortably above 30% (Figure 6) should be made an urgent priority, the most immediate and direct conclusion of this simulation analysis is that contact tracing and isolation should be viewed as an invaluable supplement to all the difficult hygiene measures and behavioural restrictions that have proven so effective^17-19^ but not as a substitute for any of them^2,20-23^.

## Implications for settings and population groups with even lower rates of symptomatic manifestations, self-reporting and confirmations

It is also important to be mindful of the fact that even the low detection and confirmation rates assumed as the default for simulations in figures 1, 3 and 5 (*θ_c_* = 0.21)^8^ may lean on the optimistic side, even in situations where transmission is broadly distributed across all age groups.^9-11^ Furthermore, even lower values may be more representative of contexts where surveillance systems are weaker and/or transmission is dominated by younger age groups, who are less likely to experience overt symptoms and correspondingly less likely to self-report for diagnosis and care. Under such conditions, even very wide transmission chains will escape detection often enough to undermine containment with contact tracing and isolation (Figure 6), making residual, uncontained, rapidly expanding transmission chains similar to figure 4 the rule rather than the exception. Particular consideration of this phenomenon is merited for contexts in which modest numbers of young people with low vulnerability from different households interact and may act as covert transmission hubs that disperse infection untraceably across a community. While large gatherings resulting in large clusters are more obvious and easier to trace, smaller pods of students in secondary schools, contact team sports and medium-sized social gatherings of youths may be just as important because SARS-CoV-2 may often quietly transit through them into several different households, workplaces and other settings without being identifiable as the focal transmission hub that connects them.

This analysis also underlines the importance of understanding the big differences that local effective reproductive numbers may have upon the traceabilitv of SARS-CoV-2 infectons, and the implications of those detection biases for interpretation of surveillance data. Moving from left to right across figure 2B, it is intuitive that much higher fractions of all cases will be detected in larger clusters arising where local transmission potential is elevated. While the attention of epidemiologists and the public alike are often captivated by large numbers associated cases arising rapidly within traceable clusters, smaller numbers of isolated cases are likely to be heavily under-represented (Figure 2B). Reported cases of community transmission may therefore represent the mere tip of a much larger iceberg that facilitates largely unnoticed, slower but more generalized spread across entire populations. Correspondingly, studies based exclusively on data from clusters of two or more associated cases^23-25^ may not be representative of transmission dynamics across entire populations and an understanding of those inherent biases should inform interpretation.

## The invaluable role of contact tracing and isolation despite the limitations imposed by covertly extended transmission chains

Having said all of the above, contact tracing and isolation should remain a vital component of any SARS-CoV-2 response. Despite the limited containment effectiveness that may be expected from contact tracing and isolation against SARS-CoV-2, it may nevertheless make an invaluable contribution to integrated suppression efforts, especially for attenuating the further spread of large outbreaks that occur at higher reproductive numbers and would otherwise facilitate much larger amounts of onward community transmission (Figure 4). This remains an invaluable and quantitatively important role for contact tracing and isolation at all stages of epidemic containment programmes, especially in relation to contexts that deliver absolutely essential services under conditions that are inherently predisposed to facilitate large outbreaks, such as hospitals, residential care homes and some factories. Furthermore, contact tracing and isolation performs an invaluable surveillance and operational research function, by providing response programmes with insights into ongoing epidemiological patterns, including the strengths and weaknesses of other interventions that could not be obtained in any other way.

## Conclusions

If indeed contact tracing and isolation is far less effective against slow, steady, undetected community transmission of SARS-CoV-2, it follows that containment and elimination efforts will have to rely predominantly upon the more burdensome and presumptive population-wide prevention measures that have proven so effective thus far.^9^ Furthermore, these will have to be sustained at a much more stringent level and for longer periods after the last detected case of community transmission than was necessary for SARS-CoV-1. Given the lack of alternatives at present, the most important missing ingredients now required for countries to progress towards elimination and exclusion of SARS-CoV-2 transmission are ambition, political will, public support, persistence and patience.

## Data Availability

All simulations provided in a supplementary file

## Contributors

GFK is the sole contributor to this article.

## Declaration of interests

I declare no competing interests.

## Acknowledgments

The author is supported by an AXA Research Chair award provided to University College Cork by the AXA Research Fund.

## Notes

### Competing Interest Statement

The authors have declared no competing interest.

## Literature cited

1. Scudellari M. How the pandemic might play out in 2021 and beyond. Nature 2020; 584: 22–5.

2. Wilder-Smith A, Chiew CJ, Lee VJ. Can we contain the COVID-19 outbreak with the same measures as for SARS? Lancet Infect Dis 2020; 20: e102–e7.

3. Smith R. Did we eradicate SARS? Lessons learned and the way forward. Am J Biomed Sci Res 2019; 6: 2.

4. Chan-Yeung M, Xu RH. SARS: epidemiology. Respirology 2003; 8 Suppl: S9–14.

5. World Health Organization. SARS (Severe Acute Respiratory Syndrome). 2020. https://www.who.int/ith/diseases/sars/en/.

6. Fineberg HV. Ten Weeks to Crush the Curve. N Engl J Med 2020; 382: e37.

7. Gandhi M, Yokoe DS, Havlir DV. Asymptomatic Transmission, the Achilles' Heel of Current Strategies to Control Covid-19. N Engl J Med 2020; 382: 2158–60.

8. Killeen GF, Kiware SS. Why lockdown? Why national unity? Why global solidarity? Simplified arithmetic tools for decision-makers, health professionals, journalists and the general public to explore containment options for the 2019 novel coronavirus. Infect Dis Model 2020; 5: 442–58.

9. Hao X, Cheng S, Wu D, Wu T, Lin X, Wang C. Reconstruction of the full transmission dynamics of COVID-19 in Wuhan. Nature 2020; 584: 420–4.

10. Chow CC, Chang JC, Gerkin RC, Vattikuti S. Global prediction of unreported SARS-CoV2 infection from observed COVID-19 cases. medRxiv 2020; https://www.medrxiv.org/content/10.1101/2020.04.29.20083485v1.

11. Li R, Pei S, Chen B, et al. Substantial undocumented infection facilitates the rapid dissemination of novel coronavirus (SARS-CoV2). Science 2020; 368: 489–493.

12. Wilder-Smith A, Teleman MD, Heng BH, Earnest A, Ling AE, Leo YS. Asymptomatic SARS coronavirus infection among healthcare workers, Singapore. Emerg Infect Dis 2005; 11: 1142–5.

13. Woloshin S, Patel N, Kesselheim AS. False Negative Tests for SARS-CoV-2 Infection - Challenges and Implications. N Engl J Med 2020; 383: e38.

14. Nachega JB, Grimwood A, Mahomed H, et al. From Easing Lockdowns to Scaling-Up Community-Based COVID-19 Screening, Testing, and Contact Tracing in Africa-Shared Approaches, Innovations, and Challenges to Minimize Morbidity and Mortality. Clin Infect Dis 2020; Electronic publication ahead of print.

15. Mizumoto K, Chowell G. Transmission potential of the novel coronavirus (COVID-19) onboard the diamond Princess Cruises Ship, 2020. Infect Dis Model 2020; 5: 264–70.

16. Woolhouse MEJ, Dye C, Etard JF, et al. Heterogeneities in the transmission of infectious agents: implications for the design of control programs. Proc Natl Acad Sci USA 1997; 94: 338–42.

17. Jefferson T, Foxlee R, Del Mar C, et al. Cochrane Review: Interventions for the interruption or reduction of the spread of respiratory viruses. Evid Based Child Health 2008; 3: 951–1013.

18. Chu DK, Akl EA, Duda S, et al. Physical distancing, face masks, and eye protection to prevent person-to-person transmission of SARS-CoV-2 and COVID-19: a systematic review and meta-analysis. Lancet 2020; 395: 1973–87.

19. Xiao M, Lin L, Hodges JS, Xu C, Chu H. Double-zero-event studies matter: a re-evaluation of physical distancing, face masks, and eye protection for preventing person-to-person transmission of COVID-19 and its policy impact. MedRxiv 2020; https://www.medrxiv.org/content/10.1101/2020.08.12.20173674v1.

20. Sanche S, Lin YT, Xu C, Romero-Severson E, Hengartner N, Ke R. The Novel Coronavirus, 2019-nCoV, is Highly Contagious and More Infectious Than Initially Estimated. medRxiv 2020; https://www.medrxiv.org/content/10.1101/2020.02.07.20021154v1.

21. Cheng HY, Jian SW, Liu DP, et al. Contact Tracing Assessment of COVID-19 Transmission Dynamics in Taiwan and Risk at Different Exposure Periods Before and After Symptom Onset. JAMA Intern Med 2020; Electronic publication ahead of print.

22. Kucharski AJ, Klepac P, Conlan AJK, et al. Effectiveness of isolation, testing, contact tracing, and physical distancing on reducing transmission of SARS-CoV-2 in different settings: a mathematical modelling study. Lancet Infect Dis 2020; Electronic publication ahead of print.

23. Xu XK, Liu XF, Wu Y, et al. Reconstruction of Transmission Pairs for novel Coronavirus Disease 2019 (COVID-19) in mainland China: Estimation of Super-spreading Events, Serial Interval, and Hazard of Infection. Clin Infect Dis 2020; Electronic publication ahead of print.

24. Qian H, Miao T, Liu L, Zheng X, Luo D, Li Y. Indoor transmission of SARS-CoV-2. MedRXiv 2020; https://www.medrxiv.org/content/10.1101/2020.04.04.20053058v1.

25. Ali ST, Wang L, Lau EHY, et al. Serial interval of SARS-CoV-2 was shortened over time by nonpharmaceutical interventions. Science 2020; 369: 1106–9.

